# Mapping the African Tobacco Control Network

**DOI:** 10.1101/2022.11.18.22282513

**Authors:** Scott J. Leischow, Olalekan Ayo-Yusuf, Janet Okamoto, Mary Warner, Jenny E. Twesten, Chad Stecher, Thomas W. Valente, Mark Parascandola

## Abstract

**Background:** To understand the state of tobacco control efforts across Africa, a first-ever survey was implemented to assess the nature and activities of tobacco control stakeholders across the African continent.

**Methods:** A survey in English, Arabic, and French was made available to individuals and organizations to assess the types and scope of tobacco control efforts and experience with tobacco control programs based on FCTC articles/MPOWER components.

**Results:** There were 219 respondents from 32 African and 6 non-African countries. Research and advocacy were the most reported activities, and several organizations emerged as network nodes for connecting tobacco control efforts across multiple African countries. The most common FCTC articles/MPOWER components worked on were (W) warning about the dangers of tobacco (58%), (M) monitor tobacco use and policies (49%), and (P) protection against secondhand smoke exposure (47%). Significant between-country differences were also found on some FCTC articles/MPOWER components: (1) (R) price and tax measures [Articles 6 and 15] (F=1.57, p=0.048), (2) industry interference [Article 5.3] (F=1.62, p=0.038), and (3) economically viable alternatives to tobacco growing [Article 17] (F=1.94, p=0.007).

**Discussion:** This study found a broad and robust tobacco control network across Africa, with multiple organizations serving those networks and having overlapping collaborations. There is considerable variability in tobacco control priorities and networking, and multiple barriers were identified to expanding the network and to fostering increased tobacco control efforts. The results point to important directions for increasing collaboration across FCTC articles/MPOWER components to improve tobacco control efforts; potential research opportunities, including an analysis of tobacco industry activities, an exploration of ways to help people quit tobacco, and approaches to elevate the cost of tobacco; and a solid tobacco control network foundation on which to build. However, exploring creative approaches to increase research most relevant to specific countries and their cultural characteristics is essential.

## INTRODUCTION

In sub-Saharan Africa, the mean prevalence of current tobacco use is around 10%.[1] Although overall prevalence of tobacco use is lower in African countries than in some other parts of the world, the African region is one of only two World Health Organization (WHO) regions (along with the Eastern Mediterranean region, which includes African countries) in which tobacco use prevalence has been increasing and is projected to continue to grow.[2] This trend reflects the impact of rapid population growth, economic growth, and growing tobacco consumption among young people. At the same time, transnational tobacco companies are aggressively marketing their products on the continent and are using legal, financial, and political strategies to counter tobacco control efforts.[3,4] Although tobacco farming has been promoted by the tobacco industry and some African countries as a means to alleviate poverty and a reason to oppose tobacco control legislation, studies show that tobacco growing has many negative consequences for the health and economic well-being of farmers.[2,5,6] Moreover, the concurrent rise in non-communicable diseases resulting from acute and chronic tobacco use threatens the fiscal sustainability of African health systems.[7]

The WHO Framework Convention on Tobacco Control (FCTC) created actionable goals aimed at reducing the supply of and demand for tobacco products.[8] A total of 51 out of 54 African countries ratified the treaty.[9] The WHO MPOWER package includes six evidence-based strategies for tobacco control aimed at improving public health: (1) monitor tobacco use and policies, (2) protect people from secondhand smoke, (3) offer help to quit smoking, (4) warn about the dangers of tobacco, (5) enforce bans on tobacco advertising and promotion, and (6) raise taxes on tobacco products.[8] Together the WHO FCTC articles and MPOWER components provide a framework and a shared lexicon for developing, implementing, and categorizing tobacco control policies and programs. Furthermore, Article 5.3 of the FCTC and related guidelines provide a framework for governmental and non-governmental organizations to act in a way that protects public health policies from tobacco industry influence.

Like other public health endeavors, the implementation of tobacco control strategies are most effective when multisector, transnational tobacco control networks collaborate to advocate for the implementation of evidence-based tobacco control policies, neutralize the tobacco industry’s influence, garner funding for research and programs, and create best practices to be shared with colleagues and governments.[10–13] Successful collaboration is predicated on effective communication and committed partnerships among a network of advocates, policymakers, funders, public health practitioners, and researchers.[14] However, there is limited information on the nature of the regional tobacco control networks in general and the African regional networks in particular. The aims of this project were therefore to: (1) map the African tobacco control community; (2) investigate the characteristics of individuals, organizations, and countries in the resultant African tobacco control network; (3) identify the FCTC articles and MPOWER components on which tobacco control actors focus; and (4) identify barriers to fostering tobacco control across the African continent. The results of this study will provide valuable information to help tobacco control networks on the African continent more effectively support tobacco control efforts in the region.

## METHODS

This project was a follow-up to the 2015 Tobacco Control in the African Continent: Research to Practice pre-conference workshop at the African Organization for Research and Training in Cancer (AORTIC) Conference in Marrakech, Morocco. The goal of the workshop was to identify research and dissemination science efforts that could advance African tobacco control practices and policies in a feasible and responsible way. Building on the workshop, the network analysis was undertaken to understand the nature of and collaborations among those involved with tobacco control across Africa.

### Instrument Design

The specific research questions were drawn primarily from network-analysis studies dedicated to improving tobacco control efforts that have been previously implemented by members of the research team[15–20] and that were informed by participants of the 2015 World Conference on Tobacco or Health[20] as well as standard network research design.[21] Researchers at the Mayo Clinic, U.S. National Cancer Institute (NCI), Arizona State University, and University of Southern California developed the survey in collaboration with the WHO Regional Offices in Africa and the Eastern Mediterranean and the Center for Tobacco Control in Africa (CTCA).

The survey was translated into three languages: English, Arabic, and French. Certified translations were obtained by the Mayo Clinic and NCI for the Arabic and French translations. The first page of the survey explained, in all three languages, the goal, aims, and rationale for the study. Survey respondents were able to choose their preferred language for survey completion.

The survey included demographic questions, such as age, education, and organizational affiliation; areas of tobacco control focus and research by FCTC article; and the types and scope of the individual respondent’s tobacco control efforts and their connections to other individuals and/or organizations. Tobacco control efforts worked on and researched were categorized by MPOWER components and FCTC articles that are particularly relevant for Africa (e.g., industry interference, farming). The survey assessed the network of individuals with whom each respondent worked, elicited suggestions for research priorities, and prompted for barriers to current tobacco control efforts. A copy of the full questionnaire is available on request.

### Study Implementation

The Mayo Clinic researchers obtained institutional review board (IRB) approval and managed survey data collection. We contacted several organizations known to be active in tobacco control in Africa to obtain the names of potential respondents or to request distribution via their listservs, recruited from an existing listserv of past participants of the AORTIC Conference tobacco control workshop, and contacted corresponding authors of published research articles focused on tobacco control efforts in Africa. The solicitation specified that we were interested in surveying individuals conducting tobacco control activities in Africa.

A snowball sampling methodology was used, whereby the survey asked participants to share the email addresses of as many as five collaborators so that we could contact them to participate in the survey. All respondents had the same 3-week contact schedule of initial email solicitation, 1-week reminder email, and final reminder email. The first wave of the survey was sent to more than 400 individuals in spring 2018, and data were collected into fall 2018 to allow for saturation. Because of the survey deployment methodology, the number of individuals who received the survey is unknown, due to posts of the survey link on tobacco control listservs; thus, the research team was unable to calculate a response rate. To incentivize and encourage participation, respondents were entered into a drawing, on completion of the survey, for a 1-year membership to the Society for Research on Nicotine and Tobacco. The drawing provided as many as 50 people with 1-year memberships (valued at $3,000).

### Consent Process

This project involved no more than minimal risk to participants, was approved by the Mayo Clinic IRB, and met all requirements of the Helsinki Agreement.

### Patient and Public Involvement

We did not include the public in the design of the study, but we did include members of the public in the conduct of the study because we used a snowball approach to circulating the survey that involved sharing the survey with a broad array of individuals. Once the paper is published, we will involve members of the public in the dissemination process because we will share it with the survey participants and others—and will encourage them to share it with others.

### Analytic Methods

Two networks were constructed based on the organizations that respondents reported working with on tobacco control. First, a country-level network was mapped by creating a network link or connection between a respondent’s country of residence and each country where an organization that is listed as a collaborator on tobacco control efforts is located. Each reported collaboration was given an equal weight, and the number of reported collaborations between two countries was used as a measure of network connection strength between the countries. This country-level network was then visualized using the Davidson-Harel layout algorithm in the “igraph” package in R, where network links between countries were given an equal weight, and network nodes (i.e., countries) were sized by the country’s number of collaborations with other countries (i.e., network degree).

The second network was constructed at the organization level based on the name of the organizations listed as collaborators on tobacco control. Similar to the country-level network, a network link was formed between a respondent’s organization and each organization that was listed as a collaborator on tobacco control efforts. This organization-level network was also visualized using the Davidson-Harel layout algorithm in the “igraph” package in R, and network nodes (i.e., organizations) were sized by the organization’s number of collaborations (i.e., network degree).

## RESULTS

### Respondent Demographic Data Overall

The respondents included 219 individuals from 32 countries in Africa and 6 countries outside the African continent. Respondents reported working in tobacco control for a mean of 10.5 years (min=0, max=42, stdev=7.7), with many holding master’s, medical, or doctoral degrees. Respondents were affiliated with a wide array of organization types based on their self-reported responses. The affiliations most represented were those from academic institutions (n=71, 32%), followed by non-profit organizations (n=32, 15%), governmental organizations (n=23, 10%), medical centers (n=21, 10%), voluntary organizations (n=18, 8%), advocacy groups (n=16, 7%), research institutions (n=17, 8%), other (n=12, 5%), consulting organizations (n=5, 2%), and intergovernmental organizations (n=4, 2%). The role in tobacco control most frequently reported by individual respondents was research (n=58, 33.7%), followed by advocacy (n=26, 15.1%), other (n=24, 14.0%), capacity-building (n=18, 10.5%), patient care (n=16, 9.3%), policy development (n=14, 8.1%), legal/law enforcement (n=12, 7.0%), and community-based program (n=4, 2.3%).

### Connectivity by Country

The average number of total connections for each country was 18.8 (min=1, max=83, stdev=23.2), while the average monthly connections was 5.2 (min=0, max=28, stdev=6.4). When only including African countries, the average number of between-country connections was 22.3 (min=1, max=81, stdev=23.0), while the average monthly connections was 6.3 (min=0, max=28, stdev=6.3). As indicated in Figure 1, there were 45 unique countries represented, and the top 10 countries with the most connections were the United States (n=83), Kenya (n=81), Nigeria (n=79), South Africa (n=79), Uganda (n=63), Zambia (n=57), Rwanda (n=30), Togo (n=26), Burkina Faso (n=25), and Tunisia (n=22). Other frequently cited non-African connections were to individuals or organizations in Canada (n=20), Switzerland (n=17), United Kingdom (n=12), Jordon (n=4), and France (n=3). In addition, there were multiple countries with one or two connections.

**Figure 1.**
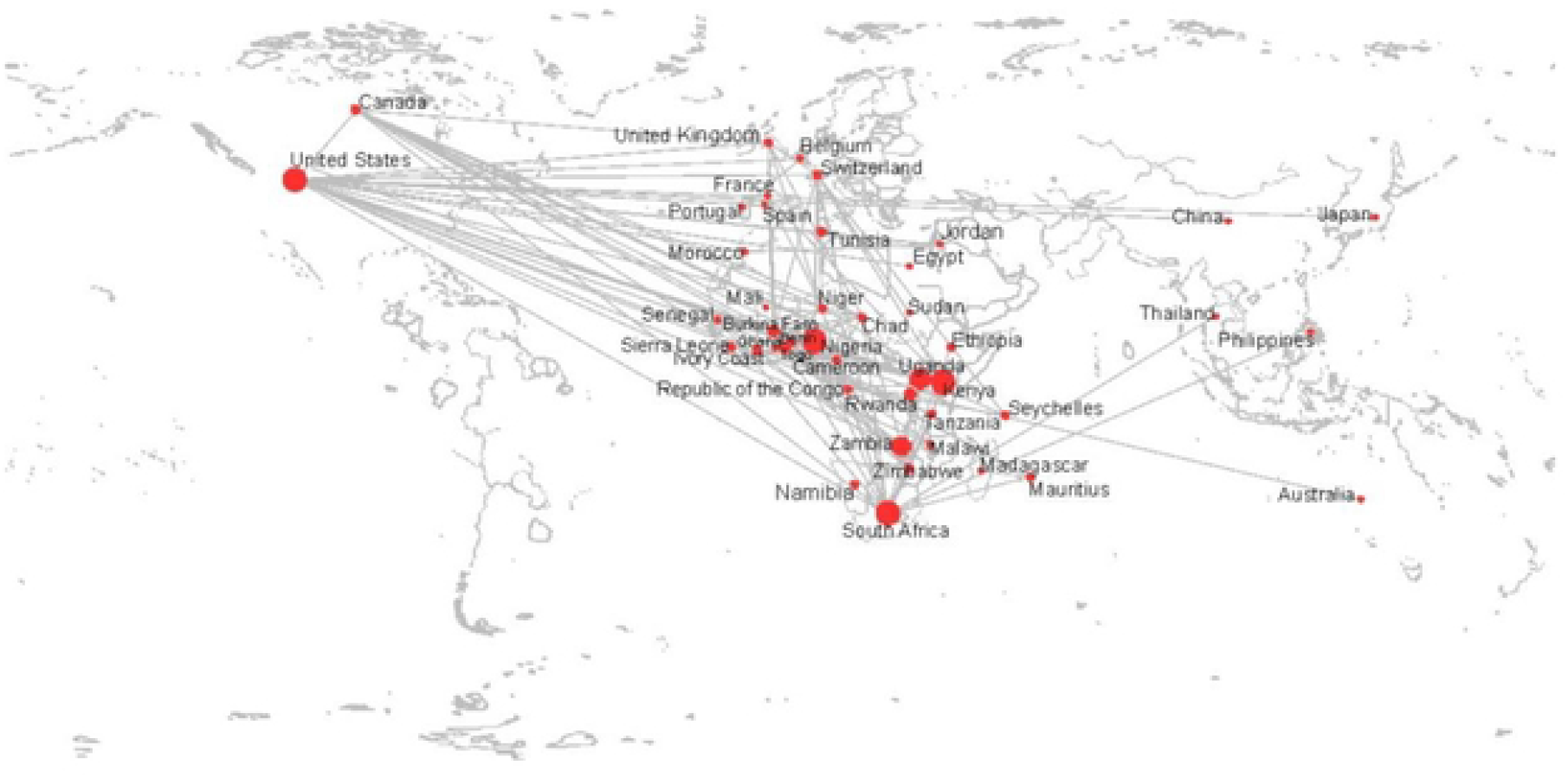
Map of Country-Level Network Connections of Collaborations on Tobacco Control in Africa

### Collaborating Organizations

As indicated in figure 2, the most commonly linked organizations in Africa included the African Tobacco Control Alliance (ATCA) (based in Togo, n=32), Center for Tobacco Control in Africa (based in Uganda, n=18), African Capacity Building Foundation (based in Zimbabwe, n=14), and International Institute for Legislative Affairs (based in Kenya, n=11). The most frequent collaborations involving organizations outside of Africa included the WHO (n=31), Campaign for Tobacco-Free Kids (n=26), and Framework Convention Alliance (n=15).

**Figure 2.**
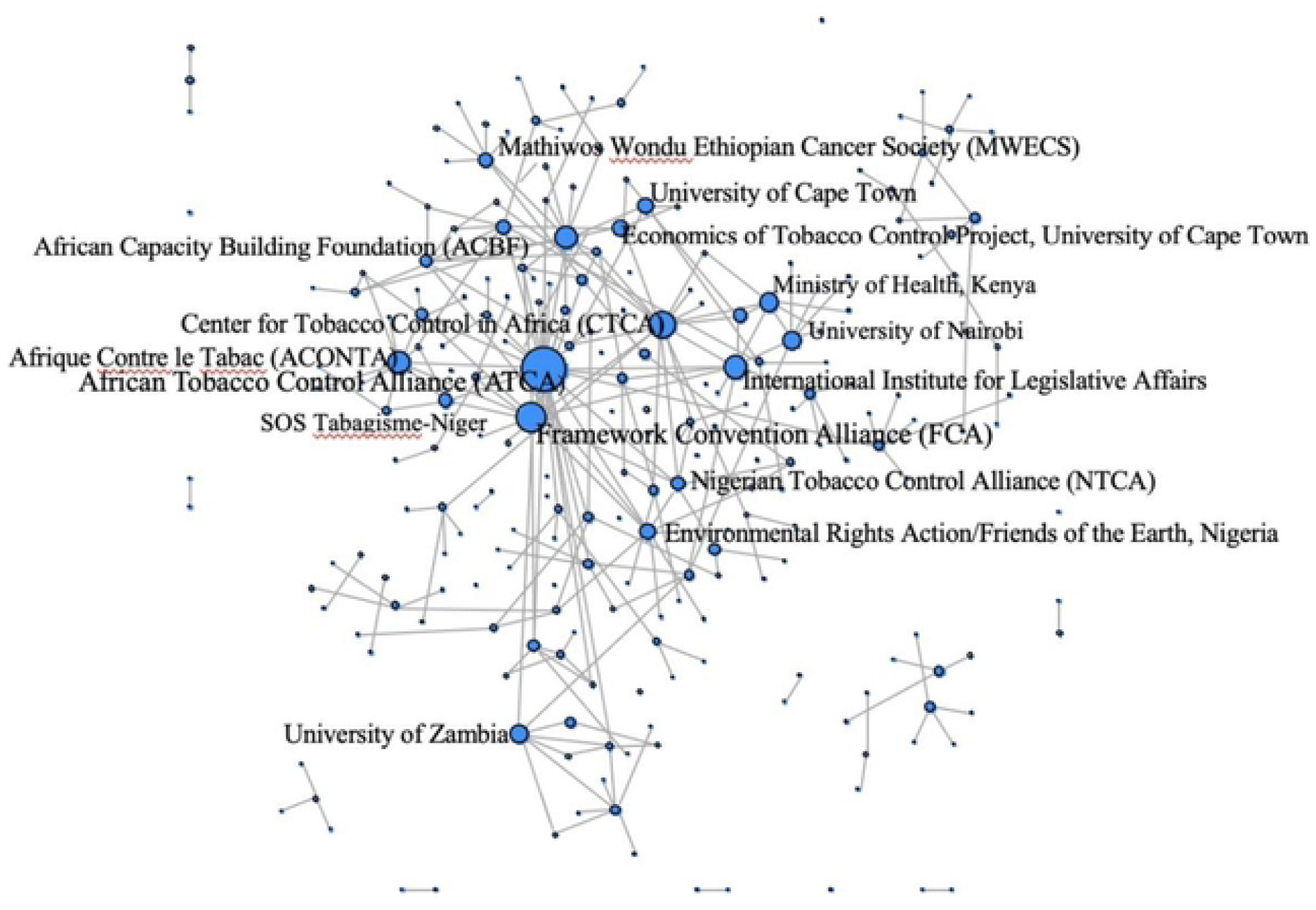
Organizations Engaged in Tobacco Control in Africa *Note:* Nodes are sized on in-degree or the number of incoming ties.

### FCTC Articles/MPOWER Components

We asked respondents which FCTC articles/MPOWER components were the focus of their work and/or research. As indicated in Figure 3, the FCTC articles most commonly worked on were (1) warn about the dangers of tobacco (W, Articles 11 and 12) (58%), (2) monitor tobacco use and policies (M, Article 20) (49%), and (3) protect against secondhand smoke exposure (P, Article 8) (47%). These top three FCTC article categories aligned with those who reported conducting research as well. (See Figure 3.) “Other” articles worked on included Articles 5.3, 9 and 10, and 16; similarly, other articles researched included Articles 5.3 and 9 and 10 and economic impacts.

**Figure 3.**
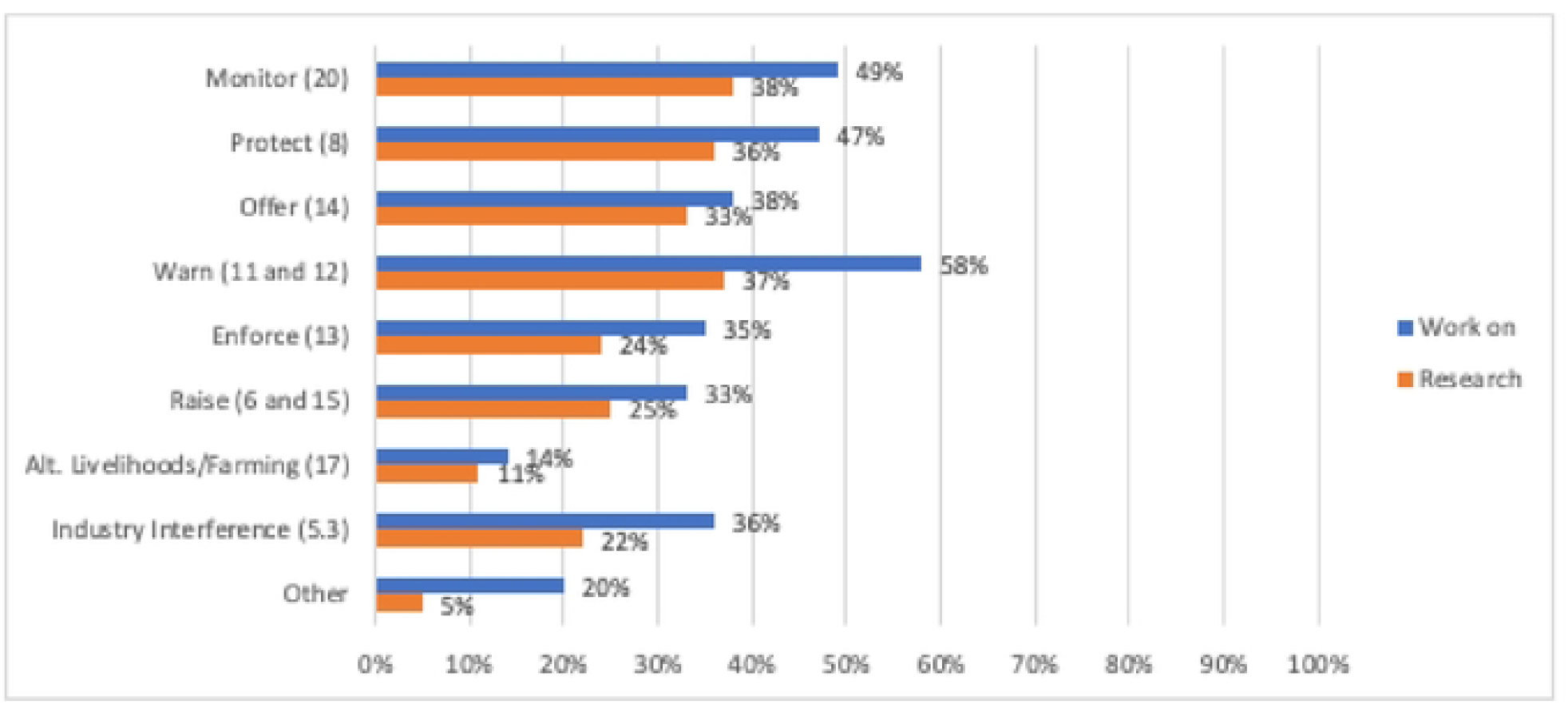
FCTC Articles and MPOWER Components Worked on and Researched by Respondents *Note:* The number to the right of the MPOWER category corresponds to the FCTC article number.

In addition, we examined whether the FCTC articles/MPOWER components worked on varied by the country in which the respondent worked. Working on certain specific FCTC articles/MPOWER components differed by country; that is, some indicated that their country worked on the following articles, and others did not: (1) raise taxes on tobacco products [R, Articles 6 and 15] (F=1.57, p=0.048), (2) industry interference [Article 5.3] (F=1.62, p=0.038), and (3) alternatives to tobacco growing [Article 17] (F=1.94, p=0.007).

The top priorities for FCTC articles/MPOWER-related research were similar across countries for some FCTC articles/MPOWER components. Respondents from different African countries indicated that the following FCTC articles/MPOWER components were important research priorities: (1) protecting people from tobacco smoke [P, Article 8], (2) warning about the dangers of tobacco [W, Articles 11 and 12], (3) enforcing bans on tobacco advertising and promotion [E, Article 13], (4) offering help to quit smoking [O, Article 14], and (5) monitoring tobacco use and policies [M, Article 20].

### Barriers to Tobacco Control in Africa

Respondents were asked to rank on a scale of 1 to 10 “the barriers you face when conducting tobacco control activities.” In order, the most commonly identified barriers to tobacco control across African countries were weak funding (mean=7.9), industry interference (mean=7.3), research not perceived as a priority (mean=6.9), lack of training (mean=6.7), lack of coordination (mean=6.2), and governmental commitment (mean=6.1).

## DISCUSSION

The goal of this study was to explore the nature and extent of collaborative relationships dedicated to tobacco control across Africa. Indeed, this first-ever analysis of tobacco control collaboration efforts across Africa demonstrates that there are extensive collaborations and that several key organizations function as network nodes. In addition, we found differences in focus on addressing specific FCTC articles/MPOWER components. The three FCTC articles/MPOWER components most commonly cited as a focus of activity were: (1) (W) warn about the dangers of tobacco use, (2) (M) monitor tobacco use and policies (epidemiology and surveillance), and (3) (P) protect from secondhand smoke exposure. Alternative livelihoods for tobacco workers (Article 17), reducing industry influence on public health policies (Article 5.3), and raising the price of tobacco (e.g., via taxes [Articles 6 and 15]) were the least common. Funding was identified as the top barrier to implementing FCTC articles/MPOWER components, followed by tobacco industry interference and the lack of a priority on research.

The fact that respondents reported limited activity around alternative livelihoods, industry interference, and tobacco taxation is surprising. Tobacco taxation is among the most cost-effective tobacco control interventions and can result in economic benefits to governments.[2,22] Yet, advancing tobacco tax policies can be challenging, as finance ministries may not prioritize tobacco control policies. Additionally, tobacco companies frequently lobby governments to oppose tobacco tax increases using erroneous arguments about illicit trade.[2] There have been successful case studies of assisting tobacco farmers to move to other crops. However, there is considerable variability in the economics of tobacco farming across countries, and some African countries, such as Malawi, are highly dependent on tobacco growing as a source of livelihoods. The challenges in moving away from tobacco dependence are complex and subject to global markets and local supply chains.[23] Further efforts in research and collaboration in tobacco control across the continent could help address the challenges in advancing tobacco price increases and in finding alternative livelihoods.

Article 5.3 was also not among the highest priority research areas identified at the time of this survey, but tobacco industry interference was ranked high as a barrier to tobacco control activities in the African region. Hence, an increased focus on activities to mitigate the risk of industry interference is essential. Recent efforts in Africa have begun to address Article 5.3. The Africa Centre for Tobacco Industry Monitoring and Policy Research, an initiative based in South Africa, has developed capacity for addressing and preventing industry interference in collaboration with partners, such as ATCA.[24] This initiative was in its infancy at the time of the survey, and a future follow-up survey would help us understand the impact of this initiative’s work.

A range of barriers were highlighted by respondents, including the lack of funding, industry interference, and the lack of prioritization and training for tobacco control efforts. Private philanthropies, the U.S. National Institutes of Health (NIH), and cancer advocacy organizations have provided some funding for specific African tobacco control and research efforts. However, preventing an increase in tobacco use in African countries will require sustained investment from more diverse sources, including from African institutions. Existing tools, such as the WHO OneHealth Costing Tool[25] and United Nations Development Program Tobacco Control Investment Case Studies,[26] could help demonstrate the return on investment of tobacco control efforts and help make the case to governments for devoting additional resources to tobacco control programs and policies.

Reducing some of the identified barriers and making tobacco control effective requires coalitions or networks that are dedicated, are data-driven, and have clear goals that can be achieved. It is clear from the results of this survey that an extensive network dedicated to tobacco control exists in Africa and is widely spread across most African countries. At the same time, as Figure 2 demonstrates, we also found considerable variability in engagement and connection. Some of those who responded to the survey had multiple ties to other tobacco control colleagues, yet many did not. This variability can be addressed and improved by organizations that function as knowledge “brokers” who can foster communication across the full network; provide training where appropriate; share new research and practice results that can benefit other members of the network; and in general, function as a hub to best ensure that the network can be optimized and can achieve a high degree of shared situational awareness.[27]

Fortunately, there are organizations that have this function, as can be seen in Figure 2. Although it appears that there are multiple highly central organizations (e.g., ATCA, CTCA), it is not clear what roles each of these central organizations is playing in the larger network. In some organizational networks, there exists a “network administrative organization” (NAO) that serves as a central knowledge broker.[19] An NAO serves as a core connector in the network, often by virtue of its funding or some administrative decision that creates a central function as the hub of the network. One follow-up of this survey that would benefit tobacco control efforts in Africa would be to characterize the roles of these centrally located organizations.

The network data make it clear that there is considerable variability in tobacco control network connectivity. Our study results show that there is an elaborate and active tobacco control network in Africa; at the same time, even greater coordination is possible. Thus, future efforts to improve tobacco control in Africa would benefit by greater strategic coordination among those central organizations to best ensure that they are linked in a way that can achieve the greatest outcomes in multiple countries. Such coordination is certainly a challenge when there are multiple funders with different priorities as well as significant variations in culture, language, resources, and governmental structures among African countries. Such variability makes increased network building and communication more challenging but at the same time more important. The tobacco industry has the resources to develop tailored approaches for each country that present a challenge for tobacco control activities to counter in the absence of effective coordination and communication among those in the tobacco control community.

However, the lack of tobacco control research as a priority and the perception of an unclear research agenda suggest that solidification of the research network could mitigate these barriers. Recent efforts to develop a research agenda and common tobacco control framework for Africa are important steps in this direction.[28] At the same time, the perceived lack of governmental commitment to tobacco control is complex and multifaceted. The lack of governmental commitment may reflect governance structures that fail to prioritize health and social well-being or to ensure policy coherence and focal points for tobacco control.[29] Furthermore, government priorities may lie elsewhere as countries address infectious diseases or other economic priorities; for example, communicable, maternal, perinatal, and nutritional conditions made up nearly 60% of lost daily adjusted life years in sub-Saharan Africa.[30]

There are multiple limitations regarding this study. First, the survey was sent to a large number of people, in part because a snowball approach was used, so we do not know how representative this sample is of those involved with tobacco control. In particular, because we included corresponding authors of research articles published over the past 20 years, we likely oversampled researchers and missed many tobacco control advocates and others who do not publish. Moreover, we do not know how many people received the request and chose not to respond, and we recognize that a snowball approach introduces bias in the sample because individuals are recommending the survey to those who they know. However, because we worked with major organizations involved with tobacco control efforts in Africa, we are confident that the results are meaningful, even if incomplete. Second, these data represent a snapshot in time in 2018 and do not necessarily reflect the current state of tobacco control efforts and collaborations. However, these data provide an important foundation for understanding the nature of the tobacco control community from the time that the survey was completed and can serve as a valuable resource to foster increased collaboration and to identify new areas of priority. Third, because this survey was conducted in 2018, we recognize that the network has changed since then. Members of our team are implementing a follow-up to this survey, so these data will serve as a baseline to assess the tobacco control network changes over time. Fourth, distinctions between institutions and activities are often not as distinct as responses might appear. For example, some respondents at academic institutions may conduct research and be activists, so the categorical responses may not always be as distinct as they would appear to be from the results.

## Data Availability

Data analyzed in this study is available via request to Dr. Scott Leischow.

## DATA AVAILABILITY

Data are available on request.

## ACKNOWLEDGMENTS

The authors would like to thank the participants of the 2015 Tobacco Control in the African Continent: Research to Practice pre-conference workshop at the AORTIC Conference in Marrakech, Morocco, for catalyzing this project. The authors are grateful to the survey respondents from the tobacco control community in Africa for their participation.

